# Exploring barriers to seeking treatment for human African trypanosomiasis due to *T.b. rhodesiense* in communities around Vwaza Marsh Wildlife Reserve in Rumphi and Mzimba Districts, northern Malawi

**DOI:** 10.1101/2023.01.04.23284208

**Authors:** Alister C. Munthali, Marshal Lemelani, Fiskani Msutu, Tisunge Zuwaki, Frederick Juma, Olaf Valverde Mordt

## Abstract

**Background:** *Rhodesiense* human African trypanosomiasis (r-HAT) remains a public health problem in Malawi, with the country reporting the highest number of cases of this acute form of sleeping sickness in the East African endemic region in 2019 and 2020. This paper explores the bottlenecks to seeking health care among patients with r-HAT around the Vwaza Marsh Wildlife Reserve (VMWR) in Rumphi and Mzimba districts in northern Malawi.

**Methodology/Principal Findings:** This qualitative study used key informant interviews (KIIs), in-depth interviews (IDIs) and focus group discussions (FGDs) to collect data. Study participants included health care workers, current and past r-HAT patients and their guardians, community leaders and traditional healers. All interviews were audio-recorded and transcribed verbatim. Content analysis was performed with NVIVO. Since r-HAT has similar signs and symptoms to malaria, patients either self-medicate or are prescribed antimalarial treatment, delaying the initiation of r-HAT treatment. While diagnosis of r-HAT can be done at health centre level, treatment requires hospitalization at the district hospital situated 60 kilometers away. Due to high levels of poverty, patients struggle to afford transport to the district hospital and their upkeep during hospitalization; patients and their guardians explained how they sold their property to fund this. In addition, patients fear undergoing very painful diagnostic lumbar puncture, which is very painful. Belief that r-HAT is caused by witchcraft still prevails, which also delays initiation of treatment. The delivery of r-HAT services is also affected by the transfer of trained health workers to other locations.

**Conclusions/Significance:** Access to r-HAT services, including treatment, could improve if community members were sensitized about the causes, signs and symptoms of the disease, and the fact that it can be effectively treated at hospital but not by traditional healers. Provision of treatment for r-HAT by health facilities around the VMWR would eliminate transport costs for patients and their guardians.

**Author Summary:** Since 2019, Malawi has reported the highest numbers of cases of rhodesiense human African trypanosomiasis. This study found that patients and their families experience many challenges in seeking health care: the disease is misdiagnosed as malaria, leading to patients receiving the wrong treatment and late initiation of the right treatment; beliefs in witchcraft cause delays as patients seek treatment from traditional healers; high levels of poverty mean that patients struggle to fund transport to the district hospital, the only facility currently providing treatment, and their upkeep during periods of hospitalization; fear of undergoing the very painful diagnostic lumbar puncture; and the transfer of well-trained health workers away from the area. There is a need to create awareness about the causes, treatment, and prevention of sleeping sickness. Once patients are diagnosed with this disease, they should be advised to seek treatment from the hospital and not from traditional healers. Patients should also be able to access treatment at the nearby health facilities rather than the district hospital, which is situated a long way from the community.

## Introduction

Human African trypanosomiasis (HAT), also known as sleeping sickness, is a vector-borne parasitic disease caused by a protozoan parasite of the genus *Trypanosoma brucei*. There are two species that cause two disease patterns in human beings: *Trypanosoma brucei gambiense* (g-HAT) and *Trypanosoma brucei rhodesiense* (r-HAT) [1]. G-HAT infection is chronic in nature, progresses slowly and causes less severe symptoms, while r-HAT infection is usually acute, causing severe symptoms and usually leading to death within a few weeks if left untreated. R-HAT is found in Southern and Eastern Africa [2] including Malawi. In 2013 the World Health Assembly adopted the WHO roadmap on neglected tropical diseases (NTDs), which, among other targets, set the elimination of HAT as a public health problem as a goal for 2020, defined as fewer than 2,000 cases of both r-HAT and g-HAT reported per year. Mainly due to national and global commitments to eliminate the disease, this target was achieved: by 2020 there were just 663 cases of HAT reported globally.

The only species found in Malawi is r-HAT, which occurs around three conservation areas, namely the Nkhota Kota Game Reserve and the Kasungu National Park (in central Malawi) and the Vwaza Marsh Wildlife Reserve (VMWR, in Rumphi and Mzimba North Districts in northern Malawi). Wild animals are the main reservoirs of r-HAT; parasites are transmitted to human beings by tsetse flies that have been infected by biting infected human beings or animals. Between 2011 and 2014, Uganda reported the highest number of cases of r-HAT of any country in Eastern and Southern Africa. However, since 2015 Malawi has been reporting the highest numbers of this disease, with the exception of 2017 when Uganda reported 13 cases and Malawi seven [3].

At a global level, there were 116 cases of r-HAT in 2019 and 98 in 2020 [3]. Table 1 shows the number of r-HAT cases reported in Malawi between 2011 and 2020.

**Table 1:** Trend in number of r-HAT cases in Malawi [3].

| Year | No. of r-HAT cases |
| --- | --- |
| 2011 | 23 |
| 2012 | 18 |
| 2013 | 35 |
| **2014** | 32 |
| **2015** | 30 |
| **2016** | 35 |
| **2017** | 7 |
| **2018** | 15 |
| **2019** | 91 |
| **2020** | 89 |

Between 2011 and 2018, Malawi reported 35 cases or less per annum, but in 2019 the number of cases increased to 91, representing 78% of the total number of cases reported globally [3]. In 2019 and 2020, 57/91 and 62/89 of the cases reported, in Malawi respectively, were from the VMWR area.

Case detection for r-HAT in Malawi is mainly passive and takes place when patients visit health facilities, with active screening done on a limited scale. Most cases of r-HAT are diagnosed at an advanced stage when symptoms are severe. Before 2013, Rumphi District Hospital was the only health facility that diagnosed r-HAT in the areas around the VMWR. Four health facilities in the area (Thunduwike, Malidade, Katowo and Mwazisi) have subsequently been upgraded and can now diagnose r-HAT. There has been a decrease in the percentage of cases resulting in death from 20.3%, between 2011 and 2013, to 8%, between 2015 and 2017 [4]. Clinical diagnosis of r-HAT is difficult because the signs and symptoms are relatively non-specific. R-HAT needs to be diagnosed and treated rapidly as the disease evolves to invade the central nervous system, eventually leading to coma and death [1]. Since initial symptoms are similar to those of malaria, it is difficult to make a differential diagnosis in resource-poor countries with limited diagnostic facilities, such as Malawi [5]. Such delays in case detection increase the risk of patients needing treatment for late-stage r-HAT, which has a fatality rate 2.5 times higher than the early stage, once the parasites cross the blood brain barrier [6].

Identified cases of r-HAT are treated with the available drugs: suramin injections for the first stage, which are associated with occasional mild reactions, and intravenous injections of melarsoprol for the advanced stage, which cause frequent and severe adverse reactions [4]. Some cases of self-cure have been reported [4]. Drugs for the treatment of sleeping sickness in Malawi are donated by the WHO through an agreement with the producers, Sanofi and the Bayer Foundation. The Ministry of Health (MoH) organizes storage and distribution to health facilities. The Foundation for Innovative New Diagnostics (FIND) helped the MoH to set up laboratory equipment in four facilities in endemic areas around the VMWR. Over the years, the Pan African Tsetse and Trypanosomiasis Campaign has supported advocacy and social mobilization [7]. The NTD programme has functional diagnosis and treatment centres in endemic districts and central reference hospitals.

R-HAT remains a public health problem in Malawi. As is the case with other endemic countries, it will be difficult for Malawi to eliminate r-HAT because of its zoonotic nature [8]. A population of infected tsetse flies is maintained among wildlife, which occasionally transmit the disease to human beings.

There is an urgent need to effectively reach at-risk populations living around the three game reserves, namely the VMWR, Kasungu National Park and Nkhota Kota Game Reserve, where this disease is endemic, and to create awareness about the aetiology, methods of prevention and diagnosis and treatment of r-HAT. Understanding people’s perceptions, beliefs and practices will help to inform policy and programming for this disease. This paper explores people’s beliefs about the causes of r-HAT and patients’ pathways for seeking health care around the VMWR in Rumphi and Mzimba North Districts, where most r-HAT cases in Malawi are detected.

## Methods

This study used an ethnographic approach to capture the lived experiences of community members seeking treatment for r-HAT. Several methods of data collection were used, including participant observation, key informant interviews (KIIs), in-depth interviews (IDIs) and focus group discussions (FGDs), as described below.

### Key informant interviews

The National Trypanosomiasis Control Programme helped to identify key informants (KIs) involved in the management of r-HAT at both district and community levels. These KIs included the programme coordinator, health surveillance assistants (HSAs), medical assistants and community leaders, including religious and traditional leaders. Sixteen purposively selected KIs were interviewed in areas around the VMWR. The KIIs focused on the aetiology and prevention of r-HAT, how the disease is experienced and factors that inhibit or facilitate seeking of healthcare services by patients with r-HAT.

### In-depth interviews

Eighteen IDIs were conducted with purposively selected community members, including former r-HAT patients, mainly to understand their perceptions, lived experiences, and strategies for coping with the disease. Interviews were also conducted with caregivers and family members who shared the challenge of seeking care within Malawi’s health care system. The IDIs helped us to better understand people’s health-seeking behaviour and drug intake during episodes of r-HAT. The informants were recruited from communities around the VMWR with the help of HSAs.

### Focus group discussions

Six FGDs were conducted with community members and between 8 and 10 participants attended each FGD: one with men, two with women, one with caregivers/guardians of current or former r-HAT patients, one with recovered r-HAT patients and one with recently treated people. The focus of these FGDs was to gather their perceptions of r-HAT, how it is experienced and the factors that inhibit or facilitate the seeking of healthcare services by patients with r-HAT.

### Recruitment and training of research assistants

Five research assistants (RAs), fluent in Tumbuka/Chichewa, with a bachelor’s degree and not less than 2 years’ experience of collecting qualitative data were recruited and trained over 4 days. The RAs helped to conduct FGDs, IDIs and some KIIs, as well as transcription. All participants in this study were ≥18 years old.

### Data management and analysis

All the transcripts were read and re-read to determine emerging themes. The major themes identified from the wider study were (i) the aetiology of r-HAT and how it is transmitted; (ii) the local terminologies for r-HAT, including signs and symptoms of the disease; (iii) health seeking behavior, including barriers to seeking treatment; (iv) prevention of r-HAT; (v) effects of r-HAT (on individuals, families and wider community); (vi) coping strategies for individuals, families and communities, and; (vii) motivation to seeking treatment. A coding framework consisting of themes and sub-themes was developed using NVivo 12 software. All transcripts were imported into NVivo 12 and relevant texts from the transcripts were coded under each sub-theme. The NVivo software was used to organize the qualitative data using a thematic content analysis approach. In this paper only thematic areas (i)-(iii) are considered.

### Ethical considerations

This study was approved by the University of Malawi Research Ethics Committee (UNIMAREC). All study participants were assured that the information they shared would remain confidential, their identity would not be disclosed, and they were free to not answer any question. Participation was voluntary. Recorders were used during the interviews and FGDs, and permission was sought from informants before tape/digital recording. Informed consent was obtained from study participants prior to the start of the interviews.

## Results

### Signs and symptoms of sleeping sickness

In areas around the VMWR in both Rumphi and Mzimba Districts, *kaskembe* is the local term used to refer to both the tsetse fly and the disease. Most informants, including current and previous patients and their guardians, community leaders and health workers reported that patients with sleeping sickness present with signs and symptoms similar to malaria, namely fever, headache, anaemia and general body pains, especially in the early stages of the disease. In addition, most study informants added that people with *kaskembe* doze or sleep during the day while chatting.

> “… According to my experience, she felt discomfort. She had fever. Then she easily went to sleep. You are speaking to her and then you realize that you are alone. She is asleep. I used to shake her to say; ‘are you with me?’ you see. ‘I am speaking to you’ then 2 minutes she is gone. She is asleep again’ yeah it was like that. Then fever came in, little by little. And then I noted that she started speaking alone …..”, (Caregiver of a patient with sleeping sickness, Thunduwike, Mzimba).

Most people, including a teacher at Thunduwike in Mzimba, reported that malaria can be differentiated from sleeping sickness because a person who has sleeping sickness tends to sleep more often. Another symptom mentioned by most informants in this study was that patients with this disease experience malfunctioning of some body parts and consequently become disabled. They feel a lot of pain, especially in the joints, and cannot walk when the disease is in an advanced stage.

> “…I could not walk instead I was crawling like a baby because I could feel pain in all joints and there was no energy for me to walk to the extent that I could feel as if I have worked over my capacity, especially this was happening with knee joints…. ”, (recovered patient, Mwazisi, Rumphi).

Other signs and symptoms mentioned by some participants included loss of appetite, oedema, dermatitis, fatigue and loss of body weight.

### Causes of sleeping sickness

All informants in this study, including health workers and community members, were aware that people get sleeping sickness after being bitten by infected tsetse flies.

> **“**Sleeping sickness comes from animals. When tsetse flies bite these animals and suck their blood they carry it. Whenever they bite human beings they transfer that parasite into human bodies and that is how sleeping sickness is caused. *Kaskembe* [the fly] is like a general player as it feeds on both human and animal blood”, (Traditional leader, Thunduwike, Mzimba).

Study participants acknowledged that sleeping sickness is very common in their community because people live very close to the VMWR and many enter the wildlife reserve without permission to poach, fetch firewood, collect mushrooms during the rainy season, produce charcoal, wash clothes and fetch drinking water and, whilst they are there, get bitten by tsetse flies.

There were, however, a few informants who believed that r-HAT is caused by witchcraft. Some informants, especially at community level, reported that some people share this belief, saying, ‘*kuli nyanga za kaskembe”*, which means there is sleeping sickness caused by witchcraft. Some people believe that this disease is caused by witchcraft and can only be dealt with by traditional healers and so do not seek formal health care services and eventually die. One clinician had patients who, despite being diagnosed with malaria and treated, did not get better. They concluded that they had been bewitched and sought traditional medicine rather than going to a health facility.

> “I have some cases which have spent quite a long period of time going from one traditional healer to the other to seek medical attention. At the time they are coming to the hospital, the disease is far much advanced …. So, this thing also happens”, (Key informant, Rumphi)

Some people around the VMWR associate sleeping sickness with witchcraft because people with this disease sleep during the daytime: they think they do this because at night they are out bewitching people. Others say they have been bewitched and so they sleep during the day instead of doing productive work. While such beliefs persist in areas around the VMWR, some informants, including an HSA at Manolo in Mzimba, reported that these days most people know that sleeping sickness is caused by tsetse flies.

Some study informants reported that it was also possible to get this disease by consuming contaminated bush meat.

> “Sometimes you may develop sleeping sickness if you eat contaminated bush meat. *…*. This is why some people who have never stepped their feet in the game reserve still get infected.
>
> Some people don’t cook meat thoroughly so the parasites that cause sleeping sickness may still be active and infect the consumers”, (P2, FGD with men, Manolo, Mzimba).

This is why one community leader discourages community members from purchasing meat from the game reserve/poachers because it is allegedly infected with the microorganisms that cause sleeping sickness. In addition, a key informant added that they have observed that new-born babies can test positive for sleeping sickness, indicating mother-to-child transmission of the disease.

### Treatment of sleeping sickness

People suffering from sleeping sickness can either self-medicate, seek treatment from traditional healers or churches or go to the health facility for diagnosis and subsequent treatment.

### Self-medication with medicines

Study participants said that the signs and symptoms of r-HAT are similar to those of malaria. People at community level are aware of malaria treatment and so when someone with sleeping sickness presents with signs and symptoms of malaria, in some cases they will purchase drugs and self-medicate. Many previous patients narrated that this was their experience, they purchased malaria medicines when they observed that they had signs and symptoms of this disease, but they did not get cured and were later diagnosed as having r-HAT. Patients that self-medicate in the early days of r-HAT may realize that they need to be tested and treated in time or they may die because they leave it too late to seek the appropriate treatment.

### Traditional healers

People with r-HAT may not initially know that they are suffering from this disease and either self-medicate or seek treatment from traditional healers, as they may think they have been bewitched.

> **“**When I started showing the signs and symptoms, I could not figure out that it was sleeping sickness. As a result, I went to different traditional healers who also told me that I was bewitched and they gave me traditional medicine which I was taking but there was no change”, (Recovered patient, Thunduwike, Mzimba).

Many previous and current patients and their guardians sought treatment from traditional healers, for example, one former patient explained that he thought he had been bewitched because he had quarreled with someone over land. In all cases of sleeping sickness, the traditional medicines that patients took did not cure them and their condition deteriorated. This was the point at which a decision was made to go to the health centre for further diagnosis and treatment. Most participants were of the view that seeking traditional medicines for the treatment of sleeping sickness is a waste of time and financial resources.

### Seeking treatment from health facilities

Initially the diagnosis of sleeping sickness in communities around the VMWR was only performed at Rumphi District Hospital, situated about 70 kilometres from the reserve. A KI at Rumphi District Hospital reported that, in addition to Rumphi District Hospital, there are 5 other health centres around the VMWR, namely Bolero, Mwazisi, Katowo, Malidade and Thunduwike, that provide diagnostic services for sleeping sickness. Many informants in this study reported that when someone is diagnosed with sleeping sickness, they are referred to Rumphi District Hospital for further confirmatory testing and subsequent treatment. The facilities around the VMWR do not provide treatment for sleeping sickness. A clinician explained that once sleeping sickness has been confirmed, the patient is given suramin if the trypanosomes are found in the blood. However, if the trypanosomes have affected the central nervous system, the patient is given melarsoprol. In addition to these two treatments, a new drug called fexinidazole (taken orally) has recently been introduced and is being piloted in a clinical trial in Rumphi.

> “… They are doing trials or investigations of that drug at Rumphi District Hospital. Currently, there are some sleeping sickness patients who have taken this treatment and other have finished taken their dosage”, (Microscopist, Manolo, Mzimba).

Many patients talked about their experience with lumbar puncture, which clinicians explained is used to determine whether the trypanosomes have entered the central nervous system; this can only be done at Rumphi District Hospital.

### Barriers to seeking treatment for r-HAT

There are several challenges experienced by community members to seeking treatment for r-HAT.

### Late diagnosis of sleeping sickness

Many participants reported that signs and symptoms of r-HAT are the same as for malaria. When patients with r-HAT visit a health facility, they are given malaria treatment based on their symptomatology. When the situation does not improve, some decide to return to the same health centre while others go to Rumphi District Hospital where they are tested for r-HAT. Participants mentioned that some clinicians recommend that patients coming from communities around the VMWR should take a test for sleeping sickness the first time they go to health facilities, while others do not. There are, therefore, delays in diagnosing r-HAT which subsequently delay initiation of the correct treatment of the disease.

### Transportation

Until recently, diagnosis and treatment of r-HAT was only available at Rumphi District Hospital, however, diagnostic services have now been established in health centres around the VMWR. Once diagnosed with r-HAT, patients are referred to Rumphi District Hospital for further tests, including lumbar puncture, and subsequent initiation of treatment. However, due to high levels of poverty, some patients do not immediately seek treatment, as Rumphi District Hospital is situated 60-75 kilometers away from these communities.

> “…. Due to problems of transport, some patients after being diagnosed with sleeping sickness, go back to their respective homes instead of going to Rumphi District Hospital for treatment because they are poor and cannot afford to pay for their transport to the district hospital for treatment. Although they have the referral letter, they still go back home and wait until they find transport to go to Rumphi District Hospital for treatment”, (Teacher, Thunduwike, Mzimba).

Study participants reported that sleeping sickness affects poor communities. Transportation is a major challenge for people in the communities around the game reserve, which explains why most patients only travel to Rumphi District Hospital when the disease is at a very advanced stage. Many study participants reported that a patient suffering from sleeping sickness cannot travel to Rumphi District Hospital alone, he must be accompanied by at least one guardian(s). Transport must also be provided for at least one guardian, making it very expensive.

> “It was very difficult for me to know that I was suffering from sleeping sickness. In addition, it was difficult for me to travel to Rumphi District Hospital due to long distance, and I had no money to use for transportation with my guardian. Due to this, my situation worsened and the brain was mostly affected which may also lead to madness ….”, (Recovered patient, Thunduwike, Mzimba).

Informants in this study estimated that the cost of one-way transport to Rumphi is around 4.4 USD per person. Lack of money for transport to Rumphi District Hospital is one of the major factors contributing to delays in seeking care for sleeping sickness.

### The process of diagnosing sleeping sickness is painful

A health worker at one of the health centres around the wildlife reserve reported that people with r-HAT are afraid of lumbar puncture when they are referred to Rumphi District Hospital: this involves the insertion of a needle into the spinal canal to collect cerebrospinal fluid for diagnostic testing of r-HAT and to stage the disease. This was reported by many participants in this study.

> “The process of confirming the diagnosis of sleeping sickness at district level is a painful process: specimen has to be taken from the spine and once one goes to Rumphi District Hospital he or she has to undergo this process ……. My patient said that it’s a painful process because they have to bend much for the doctors to get the specimen [from spine]. It takes time also for the doctors to collect the specimen. So, the patient bends for some time and when they are done collecting the specimen, it is not easy for the patient to come back to normal state ….”, (P4, FGD with guardians of patients with sleeping sickness, Malidade, Mzimba).

A game ranger reported that community members claim health workers instruct patients to bend their back until their head reaches their legs in order to take a specimen from the spinal cord, and that health workers tie patients with rubber to avoid problems with taking the specimen.

> “We are tied and squeezed with the back bent so that the head touches the knees. Other people help in pinning the patient so that there is no movement as the injection is inserted on the back. They act as if they are killing a cow”, (P1, FGD with men, Manolo, Mzimba).
>
> When community members hear such stories, they become frighted about accessing treatment for r-HAT at the health facility, which contributes to delays in seeking treatment. Some community level participants also said that some health workers are not experienced at taking specimens from the spinal cord, have to make several attempts and thus make the patient feel more pain.

### Lack of food items

During hospitalization people with sleeping sickness are provided with food by the hospital. However, guardians must provide food for themselves. Many informants reported that patients and guardians lack the necessary food and money to sustain themselves during the period of hospitalization. Guardians of patients with sleeping sickness sometimes have to work in order to provide for the patient as well as for themselves.

> “While being a guardian, I had to leave her alone and go outside and source some piece works. The kind of piece work was to do with farming. You know it was farming season. So that’s the only piece work I would have done. I leave her and do the piece work and the money I use to buy what she needs or she wants. I managed this life until she was discharged. I came back home without even a debt”, (Guardian of patient with sleeping sickness, Thunduwike, Mzimba).

The challenge to provide food and other items for the patient and guardian(s) is exacerbated by the long hospitalization period of, usually, up to 4 weeks.

### Belief in traditional healers

There are some people living in communities around the VMWR who still believe in witchcraft and that traditional healers can cure r-HAT. A microscopist at one health centre near the VMWR reported that people with such beliefs first visit a traditional healer before considering going to the hospital for treatment.

> **“**When I started suffering from this disease my wife and I visited the traditional healers to seek help because I was thinking that I have been bewitched due to what was happening. I told you that I was unable to walk, this on its own made me think it wasn’t normal at all, hence, the visit to traditional healer. And the traditional healer told me that I was really bewitched and gave me some traditional medicine”, (Patient with sleeping sickness, Thunduwike, Mzimba).

This informant said that he visited more than two traditional healers but that after receiving traditional medicines the situation did not change, which made him think of going to hospital. In some cases when a person has signs and symptoms of sleeping sickness, they and their relatives may think that they need to perform the vimbuza, a traditional healing dance popular among the Tumbuka people living in northern Malawi.

> “… I can be sick and showing many signs but some people can say its vimbuza and the person has to go to the traditional healer and dance for the evil spirits to come out of the patient. We lose a lot of lives seeking traditional medicine while that person is suffering from sleeping sickness”, (P3, FGD with women, Katowo, Rumphi).

The existence of such beliefs constitutes a major barrier to seeking treatment for sleeping sickness. Some people who had sleeping sickness and were cured acknowledged that, had they sought treatment from traditional healers, they would not have survived. Participants in this study cited many cases of people who sought treatment from traditional healers and, therefore, went to health facilities very late when the disease was well advanced and died. The belief that witchcraft is a cause of r-HAT is declining as experience increasingly demonstrates the failure of traditional medicine and the effectiveness of hospital treatment. Previously, traditional healers used to accept patients suffering from sleeping sickness, but this is changing.

> “… As of now the traditional healers no longer admit people with r-HAT. They just sensitize people about the disease. They refer the person to go to the hospital for treatment and testing”, (HSA, Mwazisi, Rumphi).
>
> “You cannot go to the traditional healers to seek remedy. A lot of people they have lost their loved ones due to this philosophy. A lot of people I tell you have lost their loved ones. The time they said; ‘let’s go to hospital’ upon arrival, the patient also dies. You see”, (P3, FGD with caregivers, Malidade, Mzimba).

One patient with sleeping sickness at Mwazisi, Rumphi said that in some cases people also resort to Pentecostal churches.

### Misperceptions about testing for sleeping sickness

Workers from health centres around the VMWR or from Rumphi District Hospital visit the reserve to test staff members and people around the reserve for r-HAT. While many informants reported that large numbers of people are tested, there are some misperceptions about testing for sleeping sickness: in some FGDs, for example with women at Katowo in Rumphi, participants reported that some community members do not want to go to hospital for r-HAT screening because they relate this to testing for HIV, thinking that health workers use the same test to diagnose HIV, and that they would rather not know their status.

> “Such people are afraid of going to the hospital to get tested even when the doctors come to our camp here, such people cannot come for the test because of the perceptions they have towards testing tools. Due to this negligence, patients may seek help or start treatment when the situation has worsened which has led to loss of many lives. However, when the doctors from Rumphi District Hospital come here they explain everything that they come to test sleeping sickness only and no other diseases and that there is no connection between sleeping sickness and HIV/AIDS”, (P4, FGD with women, Katowo, Rumphi).

Although some people have the signs of sleeping sickness, it is very difficult for them to go to hospital to be tested, despite living close to the game park.

### Poor attitude of health workers

While informants in this study appreciated the assistance they get from their health workers in nearby health facilities, some of them have a poor attitude (they are rude and they do not treat patients well) which discourages patients from visiting the health facilities.

### Shortage of staff

Health centres have been equipped to conduct tests for sleeping sickness. However, both health workers and community members reported shortages of staff, with only one microscopist at a health centre and tests only being conducted on designated days of the week. In some cases, health workers trained to conduct tests for sleeping sickness are not available when patients visit the health facility, so people return to their homes without being helped. A KI at Rumphi District Hospital reported that the programme has trained some health workers on how to manage sleeping sickness, including diagnosis and treatment. However, some of these trained health workers choose to move away or are transferred elsewhere and are replaced by staff who are neither trained nor qualified.

### Abandonment of hospitalization

While most patients adhere to the current hospitalization requirement for treatment, there have been cases where patients have escaped from hospital, mainly because of the pain but also because of other challenges experienced during hospitalization.

## Discussion

This discussion has been organized around the following themes: knowledge about r-HAT, beliefs about r-HAT, misdiagnosis and the poor attitude and perceived practices of health workers.

### Knowledge about r-HAT

This paper has outlined people’s perceptions about the aetiology and treatment of r-HAT in communities around the VMWR. As has been noted in other studies [9], community members are generally aware of the signs and symptoms of r-HAT and that these are similar to those of malaria in the early stages of this disease, which complicates finding the right therapy. Frequent sleeping is a major symptom of r-HAT, and patients have an uncontrollable desire to sleep at any time [2], even during the day, as mentioned in this study. Community members are aware of the signs and symptoms of this disease. This study has found that health workers know that the disease is caused by microorganisms called trypanosomes and that these are transmitted by tsetse flies from infected animals to human beings. While community members were aware that human beings contract sleeping sickness after being bitten by tsetse flies, they did not talk about microorganisms.

### Beliefs about witchcraft and others

Community members reported that beliefs about witchcraft causing this disease prevail in communities around the VMWR and that rHAT patients may spend long periods seeking traditional medicines. However, over the years the number of people with such beliefs has declined as they have seen that traditional healers cannot heal people with this disease while health facilities can. Studies in other countries also found that people believed they were bewitched, especially in the early days of the disease [10–13].

Diseases are perceived to be caused by witchcraft when they do not respond to treatment from health facilities: in the initial stages of sleeping sickness, especially when the disease does not respond to anti-malarial treatment, a determination is made that the disease has been caused by witchcraft or other supernatural powers. This study also found that some respondents believed that r-HAT could be transmitted from mother to child or by eating bush meat contaminated by microorganisms. WHO mentions that the disease can be transmitted from mother to child and by sexual contact, but not by eating contaminated bush meat [14].

### Misdiagnosis of r-HAT

The major challenge experienced in the management of sleeping sickness is diagnosis of the disease. This disease has similar signs and symptoms to malaria and we found that community members assume it is indeed malaria and are either prescribed antimalarials or self-medicate with painkillers, antimalarials or antibiotics. In most cases, r-HAT is diagnosed very late; Madanitsa et al. [15] argues that this is due to the weak surveillance system in Malawi. The elimination of sleeping sickness may not be achieved without improved case detection and management [16].

If a disease does not respond to treatment, other causes will be suspected and this is the point at which people with sleeping sickness will resort to consulting traditional healers, believing that the disease has been caused by witchcraft. Mulenga et. al. [10] have also argued that seeking treatment from traditional practitioners demonstrates that r-HAT is a pathology interpreted as having a supernatural origin.

In terms of treating sleeping sickness, health workers, staff from the game reserve and community members reported that all cases are currently being treated at Rumphi District Hospital. While initially even diagnosis was performed at the district hospital, since 2013 health facilities around the VMWR have been equipped to provide diagnostic services for sleeping sickness. This is similar to the situation in South Sudan, where diagnosis and treatment of sleeping sickness is offered at designated places [12].

Once diagnosed with sleeping sickness, a patient is supposed to travel to Rumphi District Hospital to initiate treatment. Study participants mentioned the barriers they experience when seeking treatment for r-HAT, including lack of transport, late diagnosis of sleeping sickness, lack of food and other items during the hospitalization period and the poor attitude of health workers. Other studies have also found that patients with sleeping sickness experience challenges, such as long distances to designated facilities that offer treatment for this disease and negative attitudes of health workers [10–12]. While in South Sudan the cost of health services was cited as a major barrier to accessing sleeping sickness services [12], in Malawi these are offered free of charge.

### The fear of lumbar puncture

Many informants reported that they feared undergoing lumbar puncture and would, therefore, not go for timely treatment. Other studies have alluded to the fact that some patients might not return for follow up visits because of fear of lumbar puncture [10,11,13,17]. In a study in Kenya, just over a third of respondents reported that lumbar puncture would prevent them from going to hospital for diagnosis [18].

### Poor attitudes and practices of health workers

Some study participants reported negative attitudes of health workers as a factor that influences informants to not seek treatment for r-HAT. Developing the target population’s trust in health workers is important to increasing uptake of screening services for r-HAT; in this study, participants said that there are others who shun screening services for r-HAT because they think that during the process they could be tested for HIV, and most of them do not want to know their HIV status. Such beliefs were also reported in Kenya [18] and in the Democratic Republic of the Congo [11], where study participants believed that an HIV test was part of the r-HAT screening procedure. As argued by the WHO [19], these negative attitudes towards hospital treatment have often led to patients absconding and not completing treatment.

### Shortage of health workers

One of the challenges reported by informants and health workers was the shortage of health workers who can manage r-HAT. The Malawi Ministry of Health acknowledges that it has a serious shortage of staff [20]. Mulenga et. al. [10] have also reported that such programmes experience high staff turnover, as reported in the current study, which affects the delivery of quality HAT services.

## Conclusion

This study has shown that r-HAT is well known in the communities around the VMWR and that most community members are aware of the way r-HAT is transmitted, namely through tsetse flies. Although traditional beliefs about witchcraft being a cause of the disease exist, they are declining. While health facilities around the VMWR can diagnose r-HAT, such facilities do not currently provide treatment: all diagnosed cases are referred to the district hospital for treatment. The major challenge to accessing treatment is the cost of travel to the district hospital and subsistence during long periods of hospitalization. Access to r-HAT services, including treatment, could be improved if community members were further sensitized about the causes of the disease, its signs and symptoms and the fact that the disease can effectively be treated at the hospital but not by traditional healers. It is also critical that health facilities around the VMWR provide treatment for r-HAT, as this would eliminate transport costs for patients and their guardians.

## Data Availability

Data contain potentially identifying or sensitive information about the participants in the study. The first author may be contacted for any further requests.

## Acknowledgments

We would like thank the health team at Rumphi District Hospital for their assistance during the collection of data for this study. We extend our thanks to the Ministry of Health for their support in the conduct of the study especially during the training of research assistants. Our special thanks go to the patients, their families and communities for their participation in this study. Thanks also to Louise Burrows of DNDi for carefully editing the manuscript.

## Notes

### Competing Interest Statement

The authors have declared no competing interest.

### Funding Statement

DNDi is grateful to the following donors for funding this project: European and Developing Countries Clinical Trials Partnership (EDCTP2) programme supported by the European Union (grant HAT-r-ACC, RIA2017NCT-1846) https://www.edctp.org/ Fundação para a Ciência e a Tecnologia (FCT), Portugal https://www.fct.pt/ Swiss Agency for Development and Cooperation (SDC) https://www.eda.admin.ch/sdc UK aid, UK https://www.ukaiddirect.org/ Médecins Sans Frontières International https://www.msf.org and other private foundations and individuals. The funders had no role in study design, data collection and analysis, decision to publish, or preparation of the manuscript.

### Author Declarations

The University of Malawi Research Ethics Committee.

## References

1. World Health Organization. Sustaining the drive to overcome the global impact of neglected tropical diseases: second WHO report on neglected diseases. 2013. Available: https://apps.who.int/iris/handle/10665/77950

2. World Health Organization, WHO Expert Committee on the Control and Surveillance of Human African Trypanosomiasis. Control and surveillance of human African trypanosomiasis: report of a WHO expert committee. 2013. Available: https://apps.who.int/iris/handle/10665/95732

3. Franco JR, Cecchi G, Paone M, Diarra A, Grout L, Ebeja AK, et al. The elimination of human African trypanosomiasis: Achievements in relation to WHO road map targets for 2020. PLoS Negl Trop Dis. 2022;16: e0010047. doi:10.1371/JOURNAL.PNTD.0010047

4. Lemerani M, Jumah F, Bessell P, Biéler S, Ndung’u JM. Improved Access to Diagnostics for Rhodesian Sleeping Sickness around a Conservation Area in Malawi Results in Earlier Detection of Cases and Reduced Mortality. J Epidemiol Glob Health. 2020;10: 280–287. doi:10.2991/JEGH.K.200321.001

5. Chisi JE, Muula AS, Ngwira B, Kabuluzi S. A retrospective study of human African trypanosomiasis in three Malawian districts. Tanzan J Health Res. 2011;13: 62–68. doi:10.4314/thrb.v13i1.61014

6. Odiit M, Kansiime F, Enyaru J. Duration of symptoms and case fatality of sleeping sickness caused by Trypanosoma brucei rhodesiense in Tororo, Uganda. East Afr Med J. 1997;74: 792–795.

7. NTD Diseases Programme Government of Malawi. Malawi NTD Master Plan 2015-2020. Lilongwe; 2014. Available: https://espen.afro.who.int/system/files/content/resources/MALAWI_NTD_Master_Plan_2015_2020.pdf

8. Partnership for Increasing the Impact of Vector Control. The role of vector control in preventing and responding to Rhodesian human African trypanosomiasis (rHAT) in Malawi. 2020. Available: https://www.afidep.org/publication/the-role-of-vector-control-in-preventing-and-responding-to-rhodesian-human-african-trypanosomiasis-rhat-in-malawi/#

9. Reid H, Kibona S, Rodney A, McPherson B, Sindato C, Malele I, et al. Assessment of the burden of human African trypanosomiasis by rapid participatory appraisal in three high-risk villages in Urambo District, Northwest Tanzania. Afr Health Sci. 2012;12: 104–113. doi:10.4314/ahs.v12i2.5

10. Mulenga P, Lutumba P, Coppieters Y, Mpanya A, Mwamba-Miaka E, Luboya O, et al. Passive Screening and Diagnosis of Sleeping Sickness with New Tools in Primary Health Services: An Operational Research. Infect Dis Ther. 2019;8: 353–367. doi:10.1007/S40121-019-0253-2/FIGURES/4

11. Mpanya A, Hendrickx D, Vuna M, Kanyinda A, Lumbala C, Tshilombo V, et al. Should I Get Screened for Sleeping Sickness? A Qualitative Study in Kasai Province, Democratic Republic of Congo. PLoS Negl Trop Dis. 2012;6: e1467. doi:10.1371/JOURNAL.PNTD.0001467

12. Bukachi SA, Mumbo AA, Alak ACD, Sebit W, Rumunu J, Biéler S, et al. Knowledge, attitudes and practices about human African trypanosomiasis and their implications in designing intervention strategies for Yei county, South Sudan. PLoS Negl Trop Dis. 2018;12: e0006826. doi:10.1371/JOURNAL.PNTD.0006826

13. Neema S, Matovu E, Valverde O. An ethnographic study of local community and peripheral health centre staff perceptions and practices regarding sleeping sickness (r-HAT) to improve treatment access and extend case detection in Uganda. Geneva; 2020 [cited 14 Dec 2022]. Available: https://dndi.org/wp-content/uploads/2021/10/HATrACC-DNDi-Uganda-rHAT-Ethnographic-Study-Report-2021.pdf

14. World Health Organization. Human African trypanosomiasis (sleeping sickness). 2022 [cited 14 Dec 2022]. Available: https://www.who.int/health-topics/human-african-trypanosomiasis#tab=tab_1

15. Madanitsa M, Chisi J, Ngwira B. The epidemiology of trypanosomiasis in Rumphi district, Malawi: a ten year retrospective study. Malawi Medical Journal. 2009;21: 22–27. doi:10.4314/mmj.v21i1.10985

16. Nolna SK, Ntonè R, Mbarga NF, Mbainda S, Mutangala W, Boua B, et al. Integration of Traditional Healers in Human African Trypanosomiasis Case Finding in Central Africa: A Quasi-Experimental Study. Tropical Medicine and Infectious Disease 2020, Vol 5, Page 172. 2020;5: 172. doi:10.3390/TROPICALMED5040172

17. Mpanya A, Hendrickx D, Baloji S, Lumbala C, da Luz RI, Boelaert M, et al. From Health Advice to Taboo: Community Perspectives on the Treatment of Sleeping Sickness in the Democratic Republic of Congo, a Qualitative Study. PLoS Negl Trop Dis. 2015;9: e0003686. doi:10.1371/JOURNAL.PNTD.0003686

18. Barasa KW. Access barriers to formal health services: focus on sleeping sickness in Teso District, Western Kenya. PhD Thesis, University of Nairobi. 2012.

19. TDR World Health Organization. Report on African trypanosomiasis (sleeping sickness) 2001. Geneva; 2003. Available: https://www.who.int/tdr

20. Ministry of Health of Malawi. Mid-term evaluation of the health sector strategic plan 2017-2022. Lilongwe; 2021.

